# Explainable machine learning-based prediction of depression severity in medical students

**DOI:** 10.1101/2023.12.14.23299975

**Authors:** Dianhao Liu, Zequn Chen, Wesley J. Marrero, Nicholas C. Jacobson, Thomas Thesen

**Affiliations:** Thayer School of Engineering, Dartmouth College, Hanover, NH, USA; Center for Technology and Behavioral Health, Geisel School of Medicine, Dartmouth College, Lebanon, NH, USA; Department of Medical Education, Geisel School of Medicine, Dartmouth College, Hanover, NH, USA; Department of Computer Science, Dartmouth College, Hanover, NH, USA

**Keywords:** Depression, Prediction, Medical Students, Machine Learning

## Abstract

**Importance:** Medical students exhibit depression or depressive symptoms at a higher rate than the general population, with a potential for reduced academic performance and increased risk of suicide and physical health problems. Understanding the factors contributing to depression severity is critical for early detection and creating support systems personalized to the mental health challenges of medical students.

**Objective:** To predict and identify the key biopsychosocial factors influencing the severity of depression in medical students across the United States using explainable machine learning algorithms.

**Design:** This prognostic study is built upon survey data from the Healthy Minds Study student survey spanning the academic years of 2009 to 2021 across the US. Students’ depression severity is measured using the Patient Health Questionnaire-9 (PHQ-9).

**Setting:** All undergraduate and graduate students from the American universities participating in the Healthy Minds Study are included in our study. While our target cohort is composed of medical students only, we include undergraduate and graduate students in our population to increase the size of our data during the machine learning training process.

**Participants:** A total of 167,999 students completed the PHQ-9 inventory, including 2,174 medical students.

**Main Outcome and Measure:** Prediction accuracy is measured by the mean absolute error across a previously unseen cohort of medical students. This metric estimates the average absolute difference between the predicted and actual responses of the general medical student population.

**Results:** By testing several predictive machine learning algorithms, we found that a sequence of binary XGBoost models achieved the lowest mean absolute error amongst all interpretable algorithms. Depression severity was best predicted by factors in the following order of importance: 1. history of depressive diagnosis, 2. disordered eating behaviors, 3. current financial stress, and 4. younger age.

**Conclusions and Relevance:** By identifying predictors of student risk for developing depressive symptoms, our findings can help facilitate early identification of medical students at risk for developing depressive symptoms and providing them with early support and proactive strategies to prevent future depressive episodes.

## 1. Introduction

The mental health of medical students is a critical issue that deserves close attention within the medical profession. Systematic reviews and meta-analyses suggest that up to a quarter of medical students suffer from depression or experience depressive symptoms, a rate significantly higher compared to the general population of the same age.(Goebert et al. 2009; Smith 2014; Puthran et al. 2016) The impact of depression on medical students is substantial, affecting both their personal well-being and professional development, leading to decreased academic performance, thoughts of leaving medical school, and a heightened risk of suicide and physical health problems.(Bahreini et al. 2016; Puthran et al. 2016) Medical students may also enter their training with high levels of depressive symptoms, emphasizing the need to address this issue within the structure of medical education. Understanding the factors contributing to depression severity is essential for creating effective intervention strategies to improve medical student mental well-being including support systems, prevention strategies, and structural changes to medical education systems.(Koutsouleris et al. 2018; Kraus et al. 2023)

Machine learning (ML) methods can be used to construct predictive models distinctly designed for understanding depression in medical students. These methods build a mathematical function to model the association between outcomes (e.g., depression severity) and input factors (e.g., psycho-bio-social variables) without making any assumptions of their underlying relationship or distribution.(Jain et al. 2021) In addition, they can easily integrate nonlinear relationships and implicitly capture interactions among factors. Such traits typically allow ML methods to achieve higher predictive performance than traditional statistical methods.

Utilizing a large multi-institutional dataset, this study aims to develop an accurate ML approach to reflect the multifactorial influences impacting the intensity of depressive symptoms in medical students. Our explainable ML approach yields an understanding of the key factors influencing depression, setting the stage for customized mental health support and targeted measures to improve the well-being and educational outcomes of medical students.

## 2. Materials and methods

### 2.1 Study population

The study population for this research is built upon the Healthy Minds Study performed by the Healthy Minds Network, a nonprofit organization dedicated to improving mental health issues among post-secondary students. Our dataset encompasses the mental health and demographic information of undergraduate and graduate students from various universities in the United States, spanning the academic years 2009-2021. Within the dataset, depression scores ranging from 0 to 27 are assigned to each student, based on their responses to the Patient Health Questionnaire-9 (PHQ-9). Any samples lacking depression scores are excluded from this study.

### 2.2 Response variable and factors

Our data comprises numerous categorical variables, many of which are poorly associated with depression. Including all these variables in ML models may significantly increase the computational burden and impede the interpretation of variables’ effects. To address these concerns, we include variables in our models based on their relative importance in predicting depression. We followed the definition from the Healthy Minds Network for depression (i.e., a PHQ-9 ≥ 10) to dichotomize the PHQ-9 scores in the variable importance analysis(Kroenke et al. 2001).

For categorical input data, two commonly employed model agnostic variable importance methods are the chi-square statistic and the mutual information statistic.(Kraskov et al. 2004) In our dataset, several variables are highly correlated, rendering the chi-square method unsuitable as it assumes independence between variables. Consequently, we assess the association between each variable and depression using the mutual information criterion. The scores derived from the mutual information criterion signify the degree of information shared between each factor and a response variable. Higher scores indicate a stronger relationship between a factor and depression. We empirically choose a threshold value representing a significant leap in mutual information scores to select the variables included in our model. Variables derived from depression (e.g., “Have you ever lost interest in things you liked for two weeks?”) are excluded from this analysis.

### 2.3 Data preprocessing

As shown in Figure 1, we exclude variables (i.e., columns) with more than 60% missing data (6 columns). Variables with 50% to 60% records missing are labeled as ‘unknown’ (1 column). In the remaining variables (16 columns), we exclude records (row) with any missing value (n=316,654). A complete summary of data missingness can be found in Supplemental Table 1.

**Figure 1.**
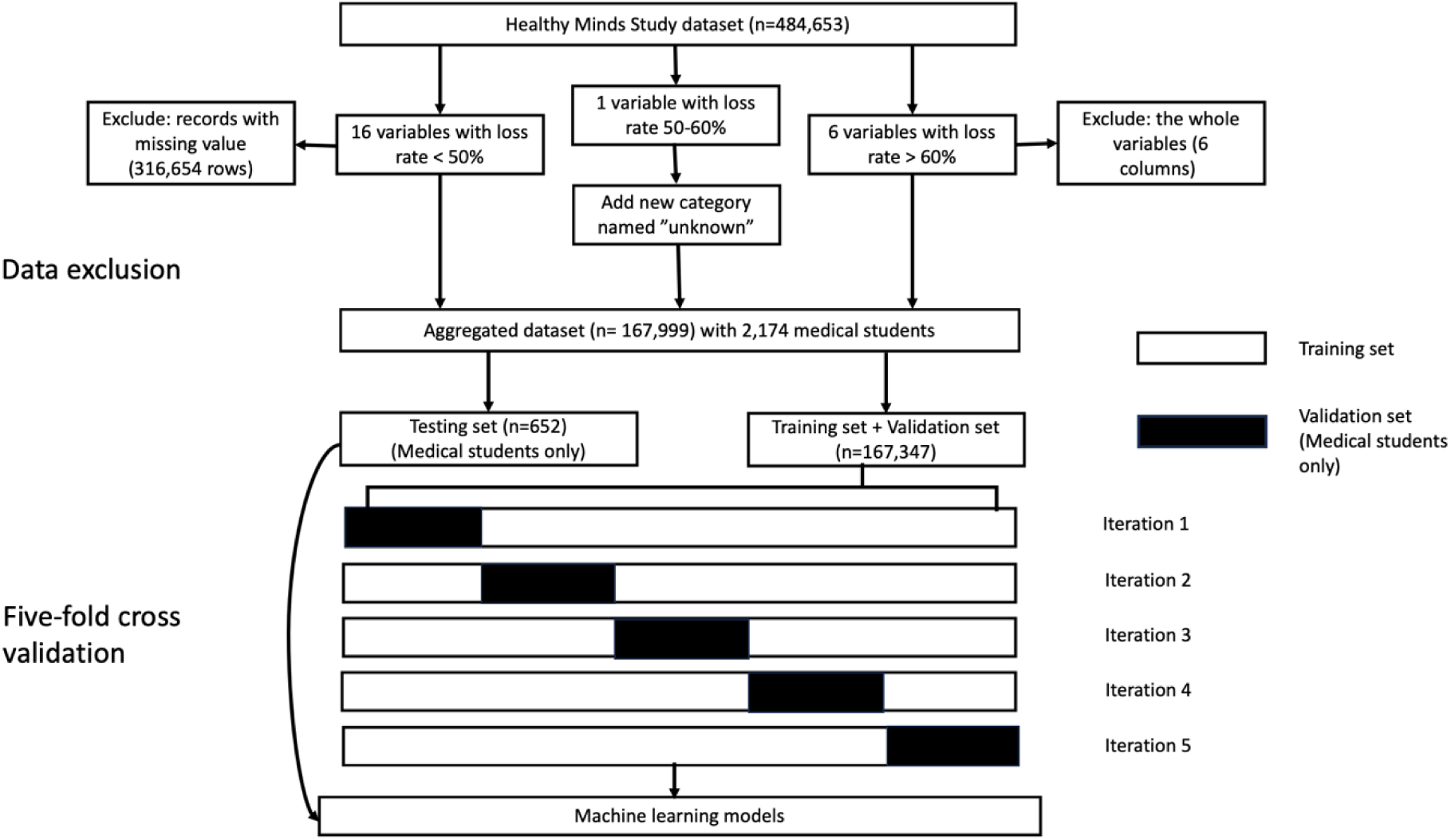
Initial data exclusion, data division, and five-fold cross validation process.

Our initial dataset consists of a small fraction of medical students and an even smaller number of medical students with elevated depression scores. Small sample sizes can affect the accuracy of ML models and prevent them from capturing complex relationships between factors and a response variable. To address this issue, we consider the enhancement of training set encompassing medical students only with other graduate students and the general (undergraduate and graduate) student population. That is, three different training set compositions are used in our ML models: medical students only, graduate students only, and the general student population. The enhanced student populations are comprised of random samples from graduate students and the general student population with the same size as the medical students’ training set. To ensure the accuracy of the prediction results in our target population, our test and validation sets only include medical students.

### 2.4 Algorithms and model evaluation

To obtain an ordinal classification model with high predictive accuracy, we employ and compare the performance of various machine learning algorithms. Specifically, we evaluate the ordinal extensions of binary classification algorithms commonly used in the mental health literature.(Cho et al. 2019; Shin et al. 2020; Lee and Kim 2022) We first consider ordinal logistic regression and support vector ordinal regression to predict the severity of depression.(Chu and Keerthi 2007) In addition, we build a multinomial neural network model to compare their results with the ordinal approaches.(Du et al. 2021) Since most machine learning algorithms have been developed for binary tasks, we convert our ordinal problem into a sequence of binary classifications.(Frank and Hall 2001) This conversion allows us to assess the performance of extreme gradient boosting (XGBoost), a widely used algorithm in the mental health literature.(Jain et al. 2021)

As shown in Figure 1, five-fold cross validation is used to prevent overfitting by tuning and evaluating the models multiple times on different training and validation datasets.(Wong and Yeh 2020) Specifically, our five-fold cross-validation is nested, which is an approach to tune and evaluate ML models within the same framework. It attempts to overcome the problem of overfitting a dataset by iterating between two layers of standard cross-validation. The inner layer involves optimizing the hyperparameters of models using a subset of the training data, while the outer layer is used to evaluate and compare the optimized models in the data not used for training (i.e., validation sets). As such, the cross-validation procedure for model hyperparameter optimization is nested inside the cross-validation procedure for model evaluation and selection. This approach ensures that our models do not excessively fit the training data during their tuning process. We assess our models using the mean absolute error (MAE), a commonly used metric representing the average absolute difference between predicted values and actual responses. Within a cross-validation framework, a model demonstrating consistently low MAE across different validation sets indicates it can be generalized to unseen data. Once the cross - validation procedure is completed, we statistically compare the MAE values of the models using Wilcoxon rank-sum tests. The best-performing explainable approach is then used to estimate the severity of depression in the previously unseen testing set. We use an overall significance level of 0.05 for our statistical analyses. The significance level and confidence interval (CI) of individual statistical tests are adjusted with the Bonferroni correction method to account for the simultaneous comparison of multiple models.

### 2.5 Factor importance and partial dependence

We use model-dependent variable importance metric to measure the relative contribution of each factor to depression severity prediction. Permutation importance is employed, involving the shuffling of single variable values while keeping the remaining factors constant.(Altmann et al. 2010) A noticeable gap in MAE between the original and shuffled input shows the shuffled factor plays an important role in the predictive model.

To get a clearer picture of how changes in factors affect the output of the best-performing model, we use partial dependence analysis.(Cafri and Bailey 2016) This analysis offers visualizations of the influence of specific factors on our model’s predictions while holding the remaining variables constant. After selecting factors with high importance, we systematically vary their values. The resulting predictions from our best-performing approach are plotted against the corresponding factor values, enabling us to observe trends and relationships.

## 3. Results

### 3.1 Descriptive results

The records after data cleaning contain information on 167,999 students, including 45,269 (26.94%) graduate students and 2,174 (1.29%) medical students. The distribution of depression levels among medical and graduate students is demonstrated in Supplemental Figure 1. Following the definition from the Healthy Minds Network for depression (i.e., a PHQ-9 ≥ 10), there are 325 (14.95%) medical students with depression. We can notice the available records shrank significantly in 2020 and 2021. This reduction in the records is due to an increasing rate of missing values during these two years, when COVID-19 was widespread. Figure 1 and Supplemental Figure 2 show the distributions of the number of overall students and students with depression by year, respectively. We select 17 variables including demographics and factors with a mutual information score above 10% (Supplemental Table 2). Supplemental Table 3 displays the demographic, physical, and mental health characteristics of our study population, including undergraduate, graduate, and medical students.

**Figure 2.**
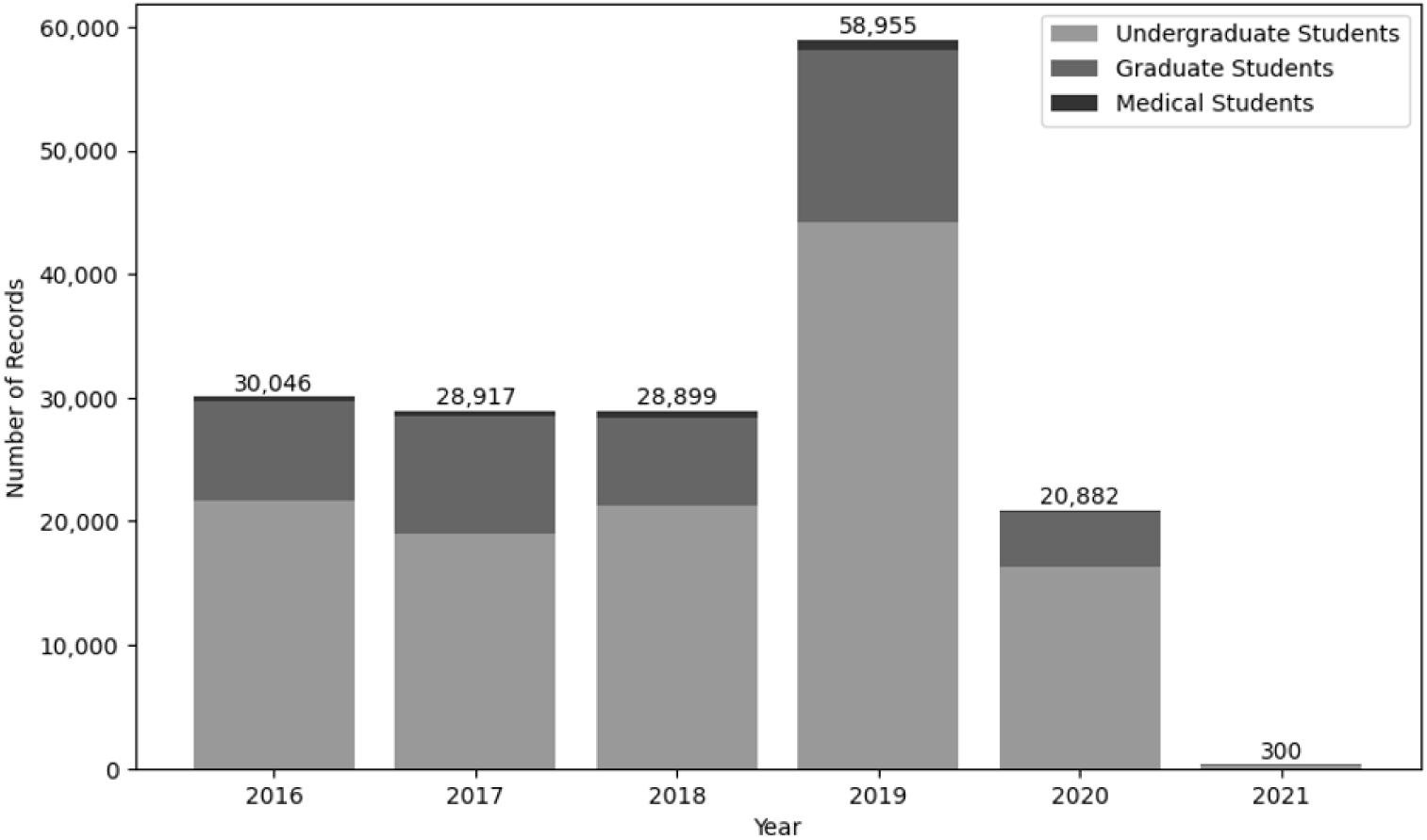
Number of records included in different years.

### 3.2 Model evaluation

The performance of our models in the cross-validation framework based on the graduate students (medical students included) training set composition is presented in Supplemental Figure 3. The remaining training set compositions are excluded for brevity as no model based on them ever outperforms any model tuned in the graduate student composition. Since we are comparing four models simultaneously at each outer cross-validation iteration and training set composition, the Bonferroni corrected significance threshold results in 0.0125 (0.05/4). We find that a neural network model significantly outperforms ordinal logistic regression (p=0.0007, 98.75% CI −0.46 to 0.46) and support vector ordinal regression (p=0.0003, 98.75% CI −0.63 to 0.63). However, the neural network is not significantly different from the sequence of binary XGBoost models (p = 0.057, 98.75% CI −0.35 to 0.35). The Wilcoxon rank-sum test across all pairs of algorithms is shown in Supplemental Table 4. When trained in the data composed of the training and validation set combined and applied to the test set, a neural network model achieves an MAE of 3.14 on a scale of 0-27, while the sequence of binary XGBoost models attains an MAE of 3.23.

### 3.3 Factor importance and model explainability

Since a neural network and the sequence of binary XGBoost models are not significantly different in terms of MAE, we evaluate the factor importance of both approaches and choose the model with better explainability. The factor importance results show the sequence of binary XGBoost achieves higher importance scores and identifies greater differences among factors than the neural network. Higher importance scores with larger differences tend to indicate that a model can better perceive the relative impact of single factors compared to the effect of interactions among multiple factors. In addition, the sequence of binary XGBoost can be represented as a tree structure, which is more intuitive than neural networks. For these reasons, we consider the sequence of binary XGBoost models as our best-performing explainable approach and use it for subsequent analyses. We show the factor importance results of the sequence of binary XGBoost models in Figure 3 and include the neural network’s variable importance in Supplemental Figure 4 for comparison purposes. Our factor importance results reveal that history of depression has the highest influence on predicted depression severity. Other notable factors include eating disorders (SCOFF [Sick, Control, One, Fat, Food] score(Morgan et al. 2000)), current financial status, and age group. Figure 4 shows the marginal impact of different levels of each of the four most important variables (i.e., variables with an importance score of at least 10%) on the predicted depression score with partial dependence plots. The rest of the partial dependence plots are shown in Supplemental Figure 5. History of depression, higher SCOFF scores, lower current financial status, and younger age are strongly associated with higher depression scores.

**Figure 3.**
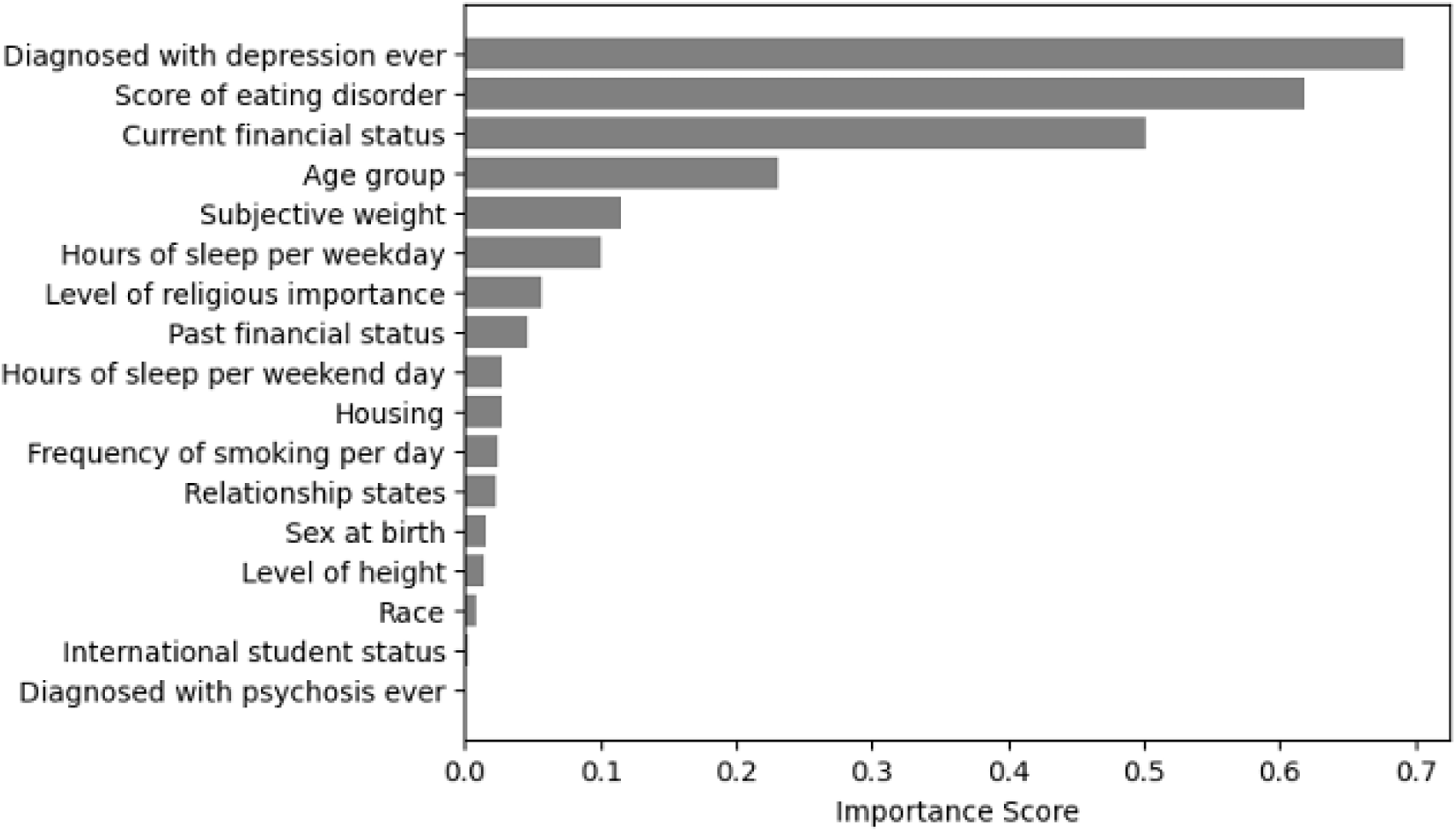
Permutation importance score in MAE of each factor in the sequence of binary XGBoost models.

**Figure 4.**
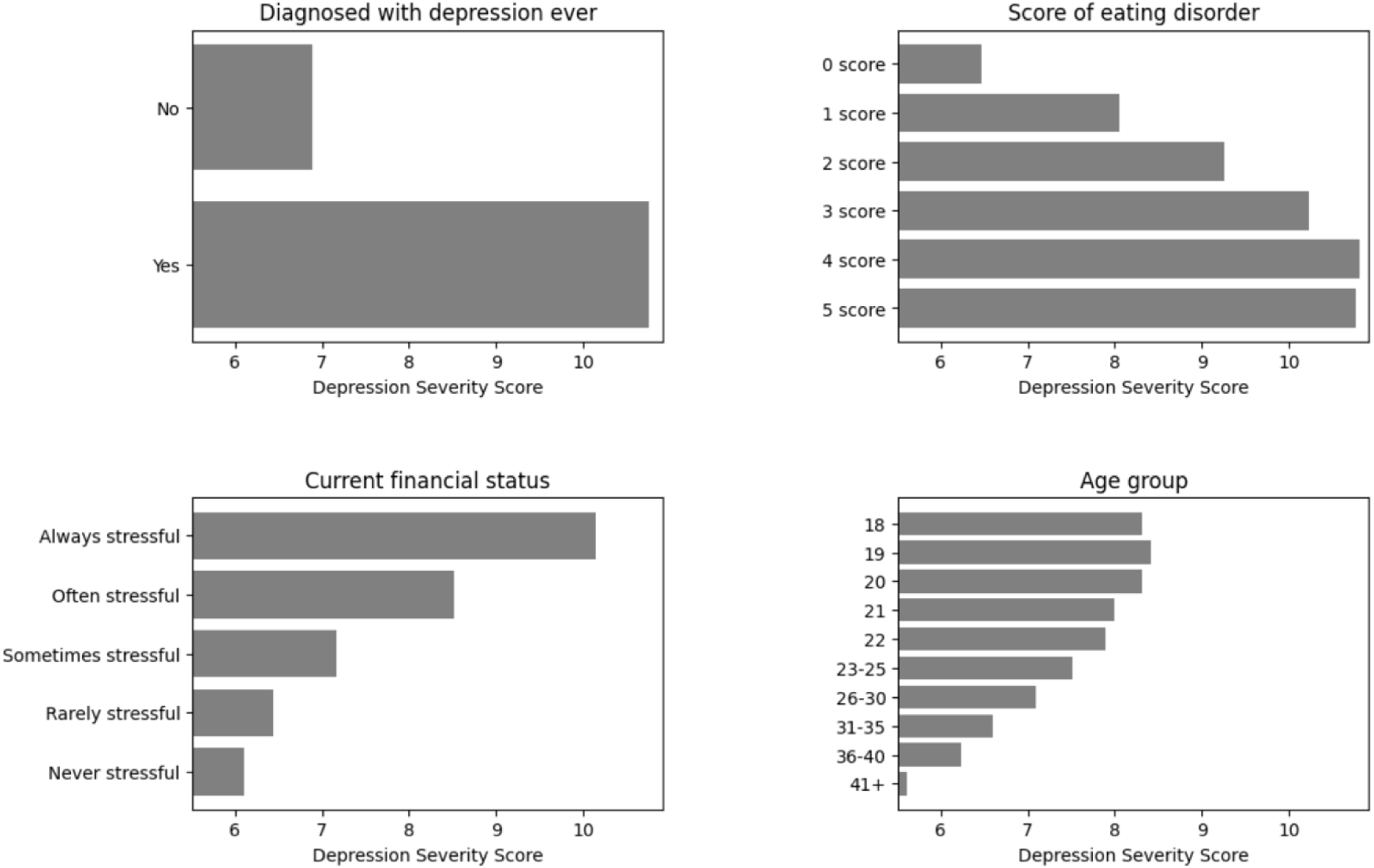
Partial dependence of depression history, eating disorder, financial status, and age group.

## 4. Discussion

In this study, the severity of depression among medical students was best predicted, in order of importance, by a history of depressive diagnosis, disordered eating behaviors, financial stress, and younger age. Less pronounced factors included perceptions of body weight deviating from normative values and diminished sleep duration during weekdays.

### Previous Diagnoses of Depression

The finding of prior diagnosis of depression as the most robust predictor reinforces the concept of depression as a recurrent disorder. Epidemiological and clinical evidence indicates that major depressive disorder generally exhibits a recurrent trajectory, with approximately one-third to one-half of patients experiencing relapse within one year after discontinuing treatment(Kessler and Bromet 2013). The likelihood of future recurrence increases with the number of previous depressive episodes(Alexopoulos et al. 1989; Gueorguieva et al. 2017). Such a pattern, coupled with the robustness of prior diagnoses as a factor in our results, underscores the need for ongoing surveillance and continuous mental health support in medical schools particularly for students with a documented history of depression.

### Disordered Eating

Disordered eating is a significant secondary predictor of depression severity. Research shows a bidirectional link between depressive disorders and disordered eating(Casper 1998). Individuals with eating disorders commonly present depressive symptoms, ranging from anhedonia to substantial feelings of worthlessness. The opposite is also documented, wherein depressive states can engender maladaptive eating behaviors due to appetite fluctuations and altered self-perception. The co-occurrence is notably prevalent in bulimia nervosa instances, with comorbidity rates spanning 31%-50%(Levinson et al. 2017). This relationship suggests the use of screening for eating disorders amongst medical students who manifest depressive symptoms or report depression histories.

### Economic Hardship

A direct relationship emerged between financial distress and heightened susceptibility to depression during medical training. Excessive tuition and ancillary educational costs contribute to financial pressures for students, especially in non-traditional students.(Destin and Svoboda 2018) These financial demands, comprising tuition, living costs, study resources, employment constraints, and potential debt accrual, represent significant stressors with implications for mental well-being. The anxiety and stress accompanying these economic strains may increase depressive symptoms.(Richardson et al. 2015) This finding suggests that educational institutions should bolster financial support mechanisms, including scholarships, and develop instructive programs addressing fiscal management.

### Age-Related Factors

Younger age emerged as a predictor of increased depression severity in medical students, consistent with prior findings that age-related developmental factors may influence mental health resilience.(Luppa et al. 2012) Older students may possess more robust coping strategies or experiential resilience, facilitating better handling of the academic and clinical demands. Conversely, younger students, possibly less experienced in navigating life’s stressors, might exhibit heightened vulnerability to stress-induced anxiety and social isolation that accompanies adjusting to the professional demands of medical school. Medical schools may consider implementing support measures tailored for younger cohorts, encompassing psychological resilience-building initiatives, mentoring, and psychologically and socially supportive environments.

### Body Perception and Sleep Patterns

The relatively weaker associations of perceived body weight deviations and reduced sleep quantity with depression, while less robust than other factors, contribute important dimensions to our understanding of depressive symptomatology among medical students. Both body image concerns and sleep disturbances are recognized in the literature as contributors to compromised mental health, especially among student populations. (Steiger and Pawlowski 2019)

### Clinical and Institutional Implications

The present findings have several implications for clinical management and institutional policies regarding student mental health. Given the identified predictors of depression severity, medical schools should employ a multifaceted approach to student mental health services, one that integrates early detection of risk factors, continuous support and monitoring of affected students, and strategies that change the structural determinants of medical student mental health. Additionally, increasing awareness about the signs of depression and related disorders like disordered eating among students and faculty could lead to earlier recognition and treatment, potentially mitigating the progression of depressive symptoms. Given the importance of sleep in well-being and learning, and the finding of lower sleep hours predicts more severe depression symptoms in this population, creating opportunities for sufficient sleep during school weeks is an important curricular change for medical schools to consider. Furthermore, with financial stress shown to be a salient risk factor, medical schools have a role to play in easing the economic burden on students. This role could involve not only the provision of financial support and education, but also advocacy for broader systemic changes that address the rising costs of medical education.

### Study Limitations

The cross-sectional design makes causal inferences about the relationships between depression severity and its factors infeasible. Longitudinal studies are required to determine the temporal progression of these associations in medical students. Additionally, the reliance on self-reported measures of depression and associated factors may be subject to biases, including recall and social desirability biases. Future studies could benefit from incorporating objective measures and more regular sampling through digital advances in passive sensing and ecological momentary assessment to enhance the validity of the findings.

## 5. Conclusion

In summary, a comprehensive approach addressing the multifaceted nature of depression among medical students is critical. This approach includes personal mental disorder history, behavioral patterns, economic conditions, and social factors that contribute to depression severity. Effective interventions require the coordination between medical educators, mental health services, financial aid departments, and policy advocates.

## Data availability

Code for analysis is available on: https://github.com/VincctsentChen/Explainable-machine-learning-based-prediction-of-depression-severity-in-medical-students-/tree/main

## Data Availability

All data produced in the present study are available upon reasonable request to the Healthy Minds Network.

https://healthymindsnetwork.org/research/data-for-researchers/

## Appendix

### Supplemental Figures

**Supplemental Figure 1.**
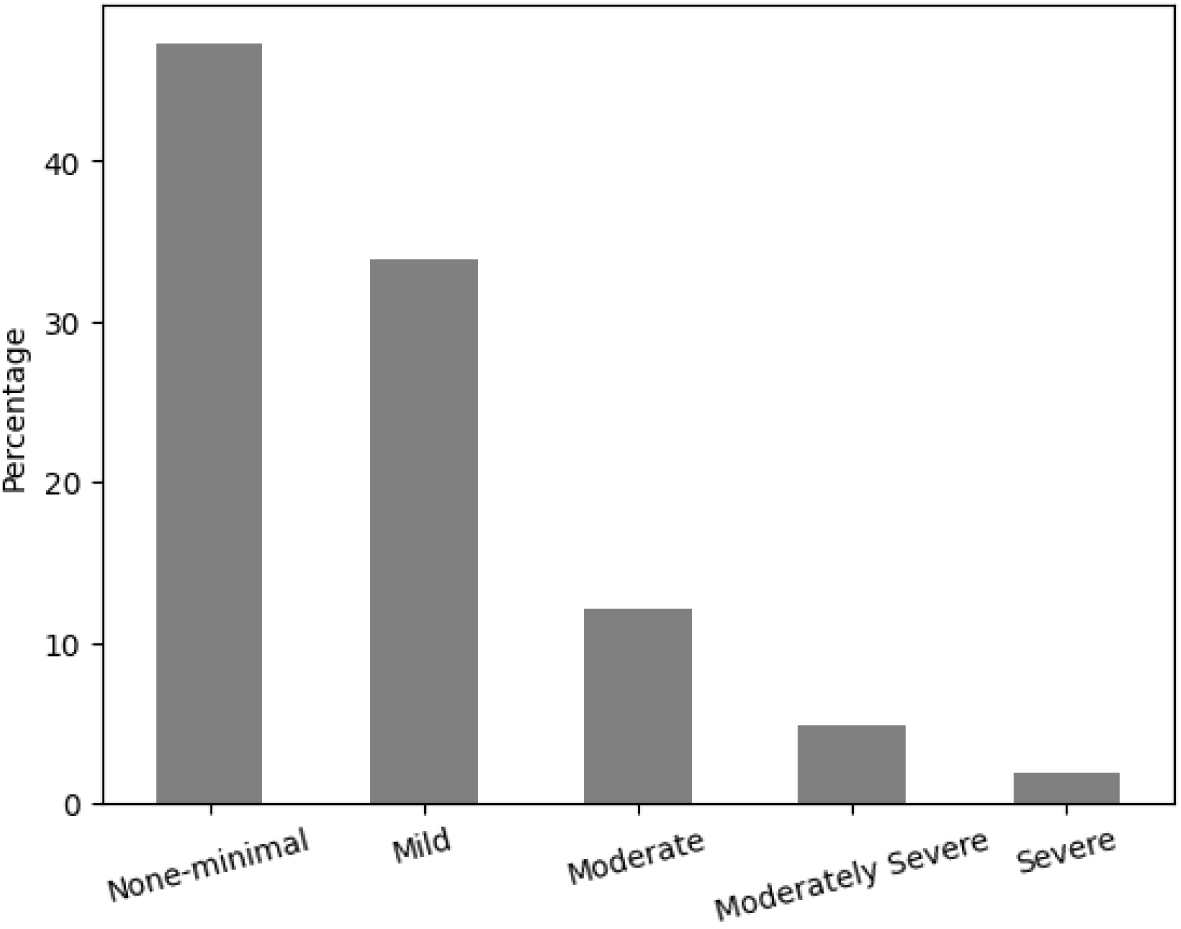
Percentage of medical students across depression categories. Definitions: PHQ-9 0-4: None-minimal; PHQ-9 5-9: Mild; PHQ-9 10-14: Moderate; PHQ-9 15-19: Moderately Severe; PHQ-9 20-27: Severe.

**Supplemental Figure 2.**
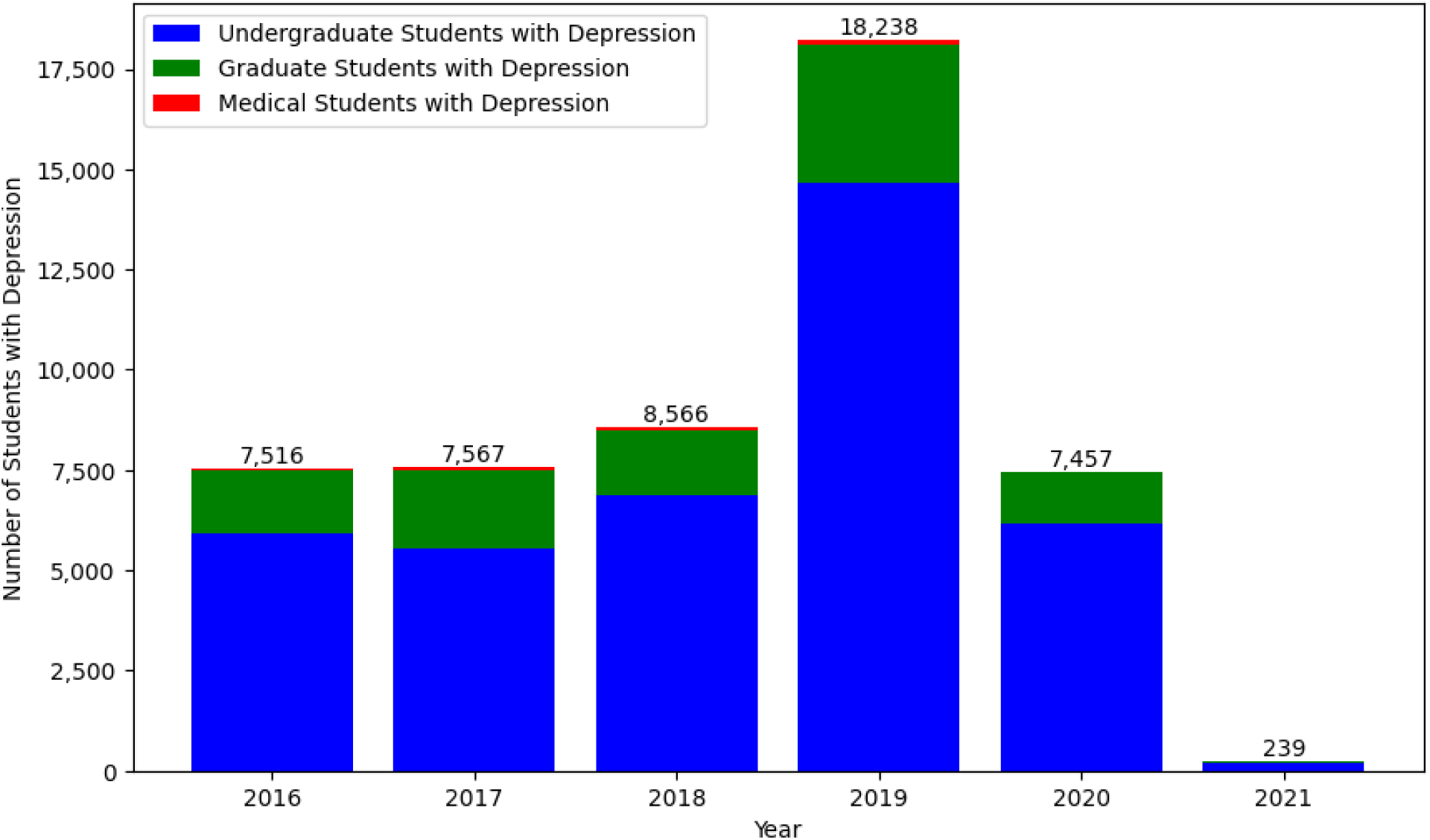
The number of students with depression across different years.

**Supplemental Figure 3.**
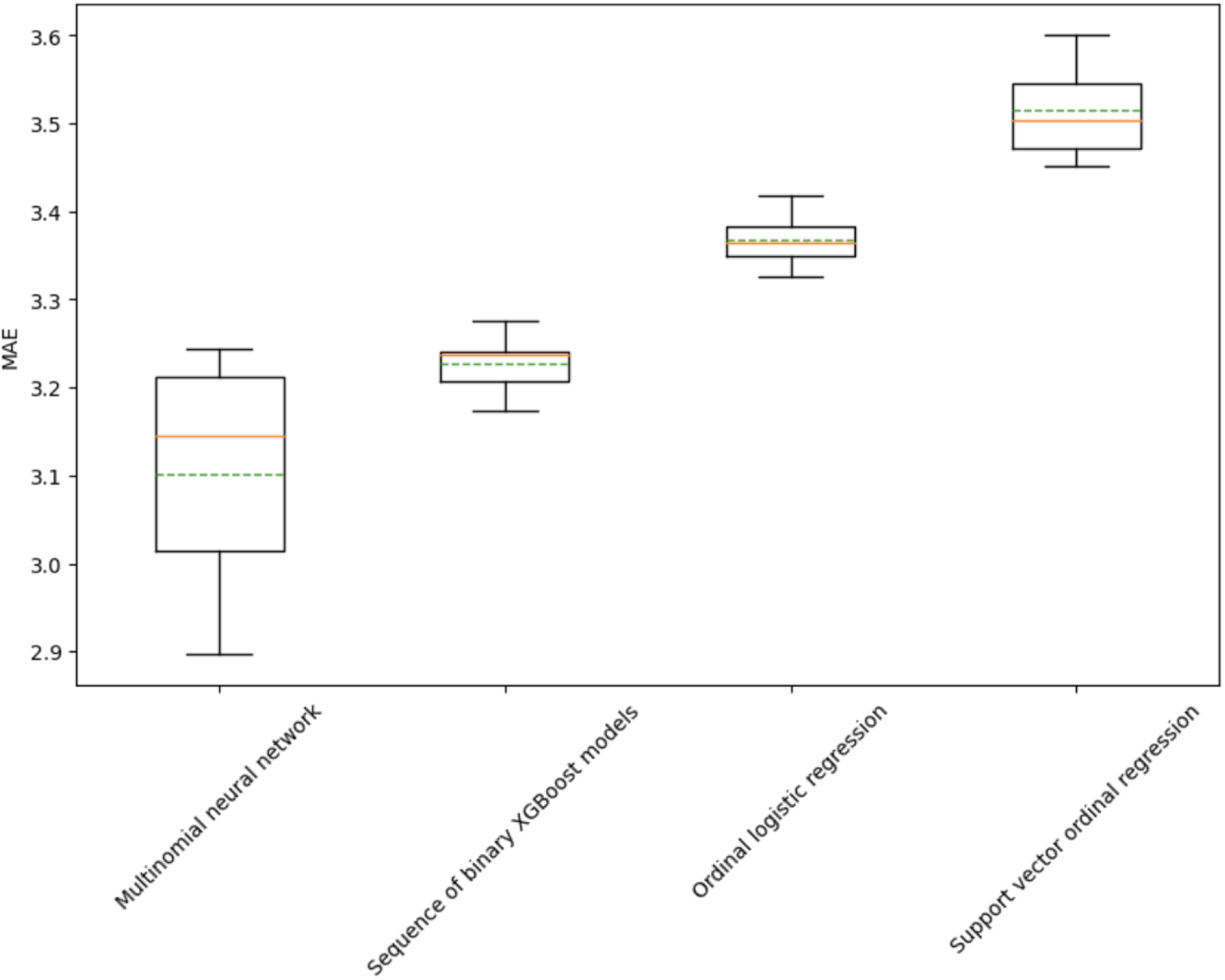
MAE of potential models to predict depression severity based on the graduate student training set composition.

**Supplemental Figure 4.**
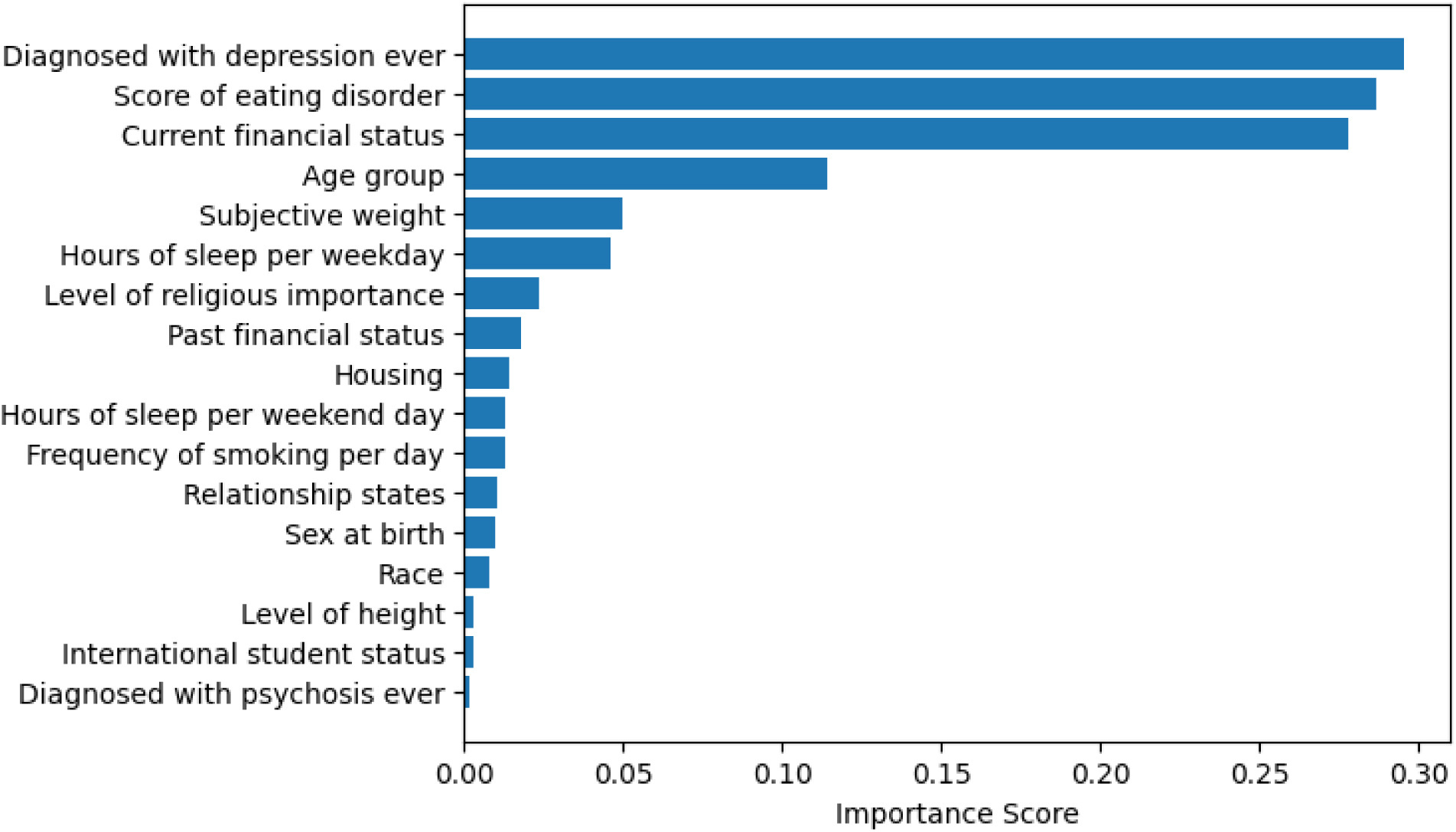
Permutation importance score in MAE of each factor in neural network model.

**Supplemental Figure 5.**
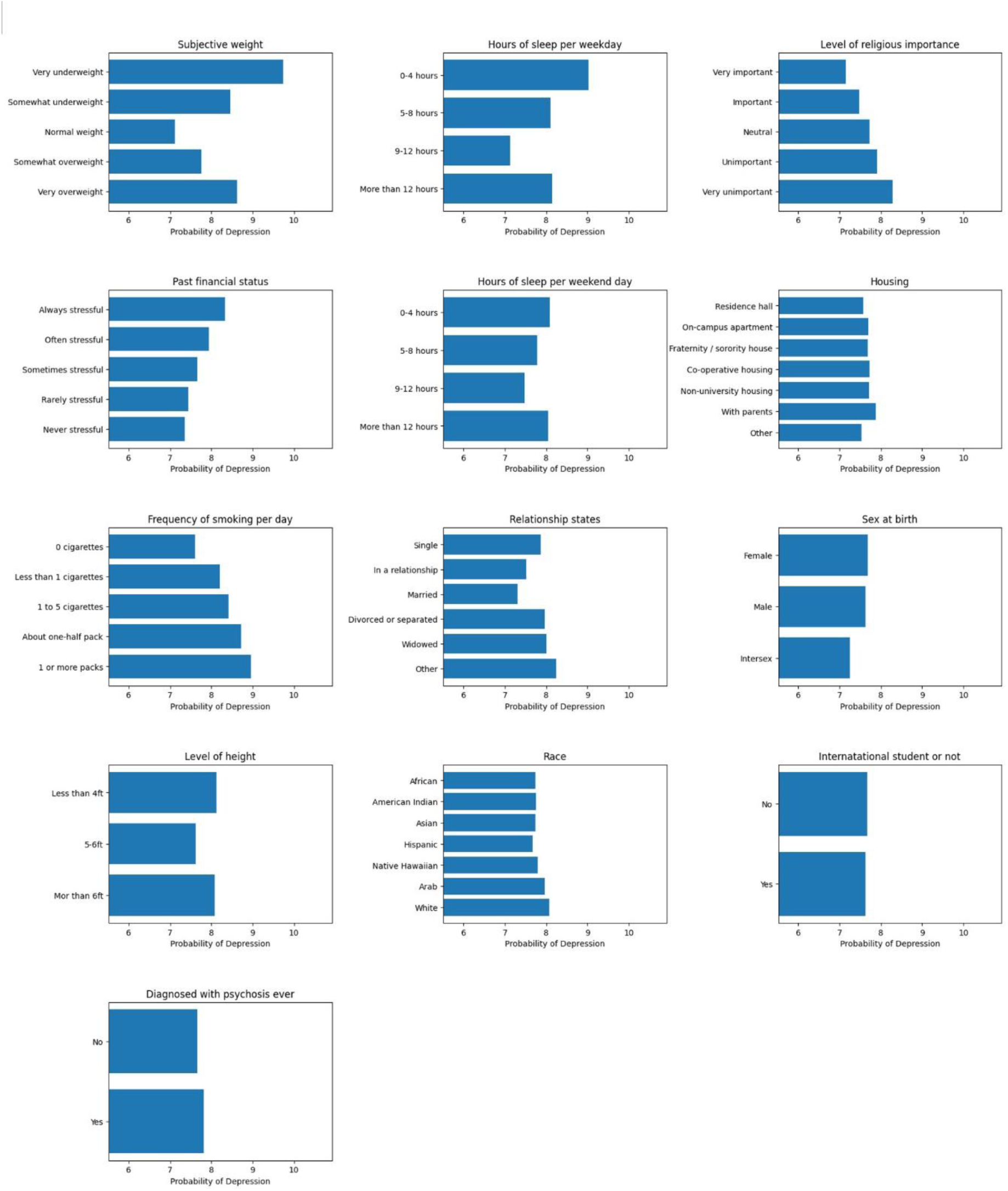
Partial dependence plot for the rest variables in the sequence of binary XGBoost models.

### Supplemental Tables

**Supplemental Table 1.** Missing data summary.

| Variable | Missing rate | Ordered factor | Levels |
| --- | --- | --- | --- |
| Eating disorder | 15.75% | Yes | 5 |
| Age group | 0.01% | No | 10 |
| Current financial status | 0.60% | Yes | 5 |
| Past financial status | 0.57% | Yes | 5 |
| Level of religious importance | 0.11% | Yes | 5 |
| International student | 6.72% | No | 2 |
| Level of height | 8.66% | Yes | 3 |
| Diagnosed with psychosis ever | 46.48% | No | 2 |
| Diagnosed with depression ever | 39.31% | No | 2 |
| Sex at birth | 0.11% | No | 3 |
| Housing | 0.09% | No | 7 |
| Race | 60.82% | No | 7 |
| Subjective weight | 21.46% | Yes | 5 |
| Relationship status | 0.07% | No | 7 |
| Frequency of smoking per day | 25.76% | Yes | 5 |
| Hours of sleep per weekday | 12.03% | Yes | 4 |
| Hours of sleep per weekend day | 12.05% | Yes | 4 |

**Supplemental Table 2.** Selected variables with their mutual information scores.

| Demographic variables |  |
| --- | --- |
| Variable | Mutual information score |
| Age group | 0.060806 |
| Sex at birth | 0.059762 |
| Race: African American | 0.068075 |
| Race: American Indian | 0.079659 |
| Race: Asian | 0.038503 |
| Race: Hispanic | 0.061001 |
| Race: Native Hawaiian | 0.062572 |
| Race: Middle Eastern | 0.029515 |
| Race: White | 0.073291 |
| International student | 0.007698 |
| Current financial status | 0.112953 |
| Relationship status | 0.006771 |
| Level of religious importance | 0.071104 |
| Level of height | 0.149735 |
| Mutual information criterion variables |  |
| Variable name | Score |
| Diagnosed with psychosis ever | 0.337642 |
| Diagnosed with depression ever | 0.245697 |
| Smoking frequency (in 30 days) level | 0.181191 |
| Have experience of being sexually assaulted ever | 0.170068 |
| Times of heavy drinking in 2 weeks | 0.163248 |
| Eating disorder (0-5 SCOFF score<sup>26</sup>) | 0.142633 |
| Sleep time on weekdays | 0.1098-0.1095 |

**Supplemental Table 3.** Demographic, physical, and mental health characteristics of the study population.

| Value | Count | Percentage |
| --- | --- | --- |
| **Degree of depression (Score)** |  |  |
| 0 | 10,765 | 6.44% |
| 1 | 9,671 | 5.79% |
| 2 | 13,648 | 8.17% |
| 3 | 14,569 | 8.72% |
| 4 | 14,034 | 8.40% |
| 5 | 12,948 | 7.75% |
| 6 | 11,972 | 7.16% |
| 7 | 11,279 | 6.75% |
| 8 | 9,920 | 5.94% |
| 9 | 8,868 | 5.31% |
| 10 | 7,196 | 4.31% |
| 11 | 6,078 | 3.64% |
| 12 | 5,520 | 3.30% |
| 13 | 4,858 | 2.91% |
| 14 | 4,115 | 2.46% |
| 15 | 3,670 | 2.20% |
| 16 | 3,251 | 1.95% |
| 17 | 2,680 | 1.60% |
| 18 | 2,446 | 1.46% |
| 19 | 1,888 | 1.13% |
| 20 | 1,570 | 0.94% |
| 21 | 1,495 | 0.89% |
| 22 | 1,168 | 0.70% |
| 23 | 958 | 0.57% |
| 24 | 920 | 0.55% |
| 25 | 623 | 0.37% |
| 26 | 374 | 0.22% |
| 27 | 643 | 0.38% |
| **Eating disorder (SCOFF Score)** |  |  |
| 0 | 88,182 | 52.76% |
| 1 | 39,463 | 23.61% |
| 2 | 23,325 | 13.96% |
| 3 | 11,641 | 6.97% |
| 4 | 3,782 | 2.26% |
| 5 | 734 | 0.44% |
| **Age group** |  |  |
| 18 | 18,707 | 11.77% |
| 19 | 23,722 | 15.30% |
| 20 | 24,103 | 15.08% |
| 21 | 23,740 | 14.55% |
| 22 | 15,220 | 9.13% |
| 23-25 | 22,620 | 12.99% |
| 26-30 | 18,842 | 10.75% |
| 31-35 | 8,184 | 4.42% |
| 36-40 | 4,485 | 2.31% |
| 41+ | 7,504 | 3.69% |
| **Diagnosed with depression ever** |  |  |
| No | 133,054 | 79.61% |
| Yes | 34,073 | 20.39% |
| **Diagnosed with psychosis ever** |  |  |
| No | 166,444 | 99.34% |
| Yes | 683 | 0.64% |
| **Current financial status** |  |  |
| Always stressful | 20,965 | 10.24% |
| Often stressful | 37,935 | 21.83% |
| Sometimes stressful | 61,138 | 41.05% |
| Rarely stressful | 35,434 | 21.49% |
| Never stressful | 11,655 | 4.79% |
| **Gender** |  |  |
| Female | 116,746 | 67.04% |
| Male | 50,345 | 32.86% |
| Intersex | 36 | 0.09% |
| **Student status** |  |  |
| Medical students | 2,174 | 1.29% |
| Other graduate students | 45,269 | 26.94% |
| Undergraduate students | 120,556 | 71.76% |
| **Level of height** |  |  |
| Less than 5ft | 10,392 | 6.22% |
| 5-6 ft | 150,835 | 90.25% |
| More than 6ft | 5,900 | 3.53% |
| **Housing** |  |  |
| On-campus housing, residence hall | 37,401 | 22.38% |
| On-campus housing, apartment | 11,111 | 6.65% |
| Fraternity or sorority house | 2,039 | 1.22% |
| On- or off-campus co-operative housing | 2,018 | 1.21% |
| Off-campus, non-university housing | 78,717 | 47.10% |
| With parents (or relatives) | 31,015 | 18.56% |
| Other | 4,826 | 2.89% |
| **International student status** |  |  |
| No | 154,575 | 92.49% |
| Yes | 12,552 | 7.51% |
| **Race** |  |  |
| Unknown | 112,902 | 61.21% |
| African | 13,981 | 7.63% |
| American Indian | 1,964 | 1.00% |
| Asian | 22,264 | 10.90% |
| Hispanic | 16,126 | 8.42% |
| Native Hawaiian | 30 | 0.05% |
| Arab | 385 | 0.29% |
| White | 20,893 | 10.50% |
| **Religious** |  |  |
| Very important | 36,294 | 14.62% |
| Important | 37,084 | 18.74% |
| Neutral | 41,240 | 26.14% |
| Unimportant | 25,578 | 21.06% |
| Very unimportant | 26,931 | 19.32% |
| **Relationship status** |  |  |
| Single | 79,323 | 47.46% |
| In a relationship | 60,842 | 36.40% |
| Married | 24,208 | 14.48% |
| Divorced or separated | 1,576 | 0.94% |
| Widowed | 184 | 0.11% |
| Other | 994 | 0.59% |
| **Frequency of smoking per day** |  |  |
| 0 cigarettes | 155,342 | 92.95% |
| Less than 1 cigarettes | 5,273 | 3.16% |
| 1 to 5 cigarettes | 3,699 | 2.21% |
| About one-half pack | 1,566 | 0.94% |
| 1 or more packs | 1,247 | 0.75% |
| **Hours of weekday sleeping** |  |  |
| 0-4 hours | 1,660 | 0.99% |
| 5-8 hours | 53,635 | 32.09% |
| 9-12 hours | 75,934 | 45.43% |
| More than 12 hours | 35,898 | 21.49% |
| **Subjective weight** |  |  |
| Very underweight | 1,461 | 0.87% |
| Somewhat underweight | 13,706 | 8.20% |
| Normal weight | 76,263 | 45.63% |
| Somewhat overweight | 60,437 | 36.16% |
| Very overweight | 15,260 | 9.13% |
| **Year that data was recorded** |  |  |
| 2016 | 29,936 | 17.91% |
| 2017 | 28,745 | 17.20% |
| 2018 | 28,650 | 17.14% |
| 2019 | 58,647 | 35.09% |
| 2020 | 20,849 | 12.47% |
| 2021 | 300 | 0.18% |

**Supplemental Table 4.** P-values and 98.75% confidence intervals on the difference of models’ performance (within parenthesis) of Wilcoxon Rank-Sum Tests. Values are shown using an upper triangular representation for simplicity purposes. P-values smaller than 0.001 are represented by dashes.

| Model | Sequence of binary XGBoost models | Ordinal logistic regression | Support vector ordinal regression | Multinomial neural network |
| --- | --- | --- | --- | --- |
| Sequence of binary XGBoost models | 1 | 0.032<br>(-0.24,0.24) | - | 0.057<br>(-0.35,0.35) |
| Ordinal logistic regression |  | 1 | 0.015<br>(-0.27,0.27) | - |
| Support vector ordinal regression |  |  | 1 | - |
| Multinomial neural network |  |  |  | 1 |

